# Similarities between long COVID and cognitive impairment and potential implications; Results from the 2022 BRFSS

**DOI:** 10.1101/2024.04.02.24305215

**Authors:** Mary L. Adams, Joseph Grandpre

**Author notes:** Corresponding author: Mary Adams, 247 N Stone St, West Suffield, CT 06093. The authors have no conflicts of interest to disclose.

## Abstract

**Background:** COVID has been linked to cognitive issues with brain fog a common complaint among adults reporting long COVID (symptoms lasting 3 or more months).

**Objective:** To study similarities and differences between cognitive impairment (CI) (the cognitive disability measure) and long COVID.

**Methods:** Using 2022 BRFSS data from 50 states and 169,894 respondents in 29 states with COVID vaccine data, respondents with CI and long COVID were compared in unadjusted analysis and logistic regression. Apparent vaccine effectiveness was compared in the 29 states.

**Results:** Prevalence of long COVID was 7.4% (95% CI 7.3-7.6) and CI was 13.4% (13.2-13.6) with both rates higher among women, ages 18-64 years, Hispanics, American Indians, ever smokers, those with depression, e-cigarette users, and those with more of the co-morbidities of diabetes, asthma, COPD, and obesity. The strong association between long COVID and CI was confirmed. Apparent vaccine effectiveness of 3 or more doses vs <3 was 38% for long COVID and 35% for CI, in both cases reducing rates for 3 or more doses to those comparable to adults with 0 comorbidities and showing dose response gradients. For CI, apparent vaccine effectiveness was similar for respondents with or without long COVID. Logistic regression confirmed most results except the magnitude of vaccine effectiveness on CI was reduced in some models while vaccine effectiveness for long COVID was confirmed.

**Conclusions:** More research is needed to understand the apparent effectiveness of COVID vaccines on CI but, if confirmed, results could expand the list of non-infectious outcomes for which mRNA vaccines can be effective.

## Background

Cognitive problems such as “brain fog” are a common complaint among adults reporting long COVID (symptoms lasting ≥3 months)(1). A recent publication (2) describes Post-COVID cognitive dysfunction (PCCD) as a condition in which patients who had long COVID exhibit subsequent cognitive impairment that cannot be explained by an alternate diagnosis. Any possible connection between COVID-19 and cognitive difficulties suggests the potential need for surveillance. One problem with surveillance of cognitive impairment is the lack of standardization in measurement (3,4). To help meet requirements of the Affordable Care Act (5) for measurement of disability, the Department of Health and Human Services published guidelines (6) that include questions to be used on federal surveys, including the Behavioral Risk Factor Surveillance System (BRFSS). One of those disability questions is “Because of a physical, mental, or emotional condition, do you have serious difficulty concentrating, remembering, or making decisions?” The question appears to be an acceptable measure for cognitive impairment but not cognitive decline because it lacks a time frame (3,4).

## Objective

The main objective was to study similarities and differences between cognitive impairment (CI) using the cognitive disability measure and long COVID. To fill in more background we opted to start with the trend data for CI from 2013 when the question was first included on the BRFSS. Another objective was to determine approximate effectiveness of COVID vaccines on reducing rates of long COVID, with other outcomes including CI used for comparison.

## Methods

The study used BRFSS data from 50 states and DC that are available on the Centers for Disease Control and Prevention (CDC) website (7) along with survey questions and information needed for analysis. Data were already weighted to adjust for the probability of selection and to reflect the adult population of each state by gender, age, race/ethnicity, education level, marital status, home ownership, regions within states, and telephone source. Data for each year from 2013 to 2022 were downloaded from the BRFSS website (7) to monitor CI rates for different age groups over that time frame. For the 2022 data, the median response rate for the 50 states plus DC for land line and cell phone surveys combined was 45.1% (7), ranging from 36.2% to 66.8%, which is typical. A total of 385,617 respondents were included for 2022 for the 50 states plus separate analyses were done on 169,894 respondents from the 29 states that asked the COVID vaccine module on 3 different survey versions which were combined per CDC instructions (8).

Measures: Cognitive impairment (CI) was ascertained from this question: “Because of a physical, mental, or emotional condition, do you have serious difficulty concentrating, remembering, or making decisions?” COVID questions on all 2022 BRFSS surveys addressed ever testing positive for COVID-19 and if so, did any symptoms last 3 months or longer, which was defined as long COVID (7). Other measures included age, race/ethnicity, gender, income, education, employment, a depression diagnosis, census region (Northeast, Midwest, South and West), e-cigarette use, and any HIV risk factor in past year (injected drug use, STD treatment, or exchanging sex for money or drugs, had anal sex without a condom, or had four or more sex partners). Obesity (body mass index ≥30 based on self-reported height and weight), sedentary lifestyle (no leisure time physical activity in the past month), current smoking, and depression (ever told they had a depressive disorder), self-reported asthma, diabetes, cardiovascular disease (CVD; heart attack, angina, coronary heart disease, or a stroke), and Chronic Obstructive

Pulmonary Disease (COPD) were also included. A composite measure of 5 chronic conditions/risk factors (obesity, asthma, CVD, diabetes, and COPD) among 6 found to be associated with US hospitalizations for COVID (9,10) was included and termed “COVID risks”. In addition, a composite measure of 5 risk factors shown to be associated with cognitive decline and dementia (11) included obesity, diabetes, depression, sedentary lifestyle, and current smoking and was termed “dementia risks”. Both sets of risk factors also included hypertension which was excluded in this study because data on hypertension was not available for 2022. A third risk measure combined these two sets of measures, but with removal of duplicates and current smoking has 7 measures (“combined risk”: the 5 COVID risks plus depression and sedentary lifestyle). Vaccine measures for the 29 states included receipt of any COVID vaccine, number of doses, and ≥3 doses vs <3 (12).

Analysis: Stata version 18.0 (StataCorp LLC, College Station TX) was used to account for the complex sample design of the BRFSS in unadjusted analysis and controlled for the listed factors in logistic regression. The survey measures used to describe the survey design were _psu and _ststr, weight=_llcpwt or the survey version weight; linearized variance estimation was selected, with the option to center at the grand mean for strata with a single sampling unit. Missing values for any measure except income were excluded from analysis. For CI trend data for 2013-2022, in addition to all ages, data for ages 18-24 years, adults <45 and ages 45+ years were tracked.

Separate univariate analysis was done for long COVID and CI from 2022 data and variables to be included in logistic regression models were selected from these results. Apparent vaccine effectiveness was determined by comparing prevalence rates for long COVID and CI respectively for each additional vaccine dose from 1 to ≥4 compared with 0 and for ≥3 vaccine doses vs <3, the latter measure including all adults in the 29 states with non-missing vaccine values (12).

## Results

Figure 1 shows the rates of CI from 2013-2022 by selected age groups indicating that rates for adults age 45+ are relatively constant while rates for ages <45 and especially those for 18-24- year-olds increased, notably after 2020. Prevalence of CI in 2022 for all ages was 13.4% (95% CI 13.2-13.6) and of long COVID was 7.4% (95% CI 7.3-7.6) representing 21.9% of all adults testing positive for COVID (12). Overall, one in 5 respondents with a positive COVID test reported long COVID, ranging from one in 7 in HI to 1 in 3.4 in WV. The CI and long COVID rates were both higher among women, ages 18-64 years, Hispanics, American Indians, ever smokers, those with depression, e-cigarette users and those reporting HIV risk factors and both measures were lowest in the Northeast region (Table 1). Rates of both long COVID and CI increased up to at least 2-fold with more of each of the 3 composite risk measures, with CI tending to increase the greater amount whether the risks were originally associated with dementia or COVID. For comparison, results for the 34.8% of adults with a positive COVID test (not shown) had relatively small changes for any of the 3 composite measures of risk. The strong association between long COVID and CI was also confirmed (Table 1).

**Figure 1.**
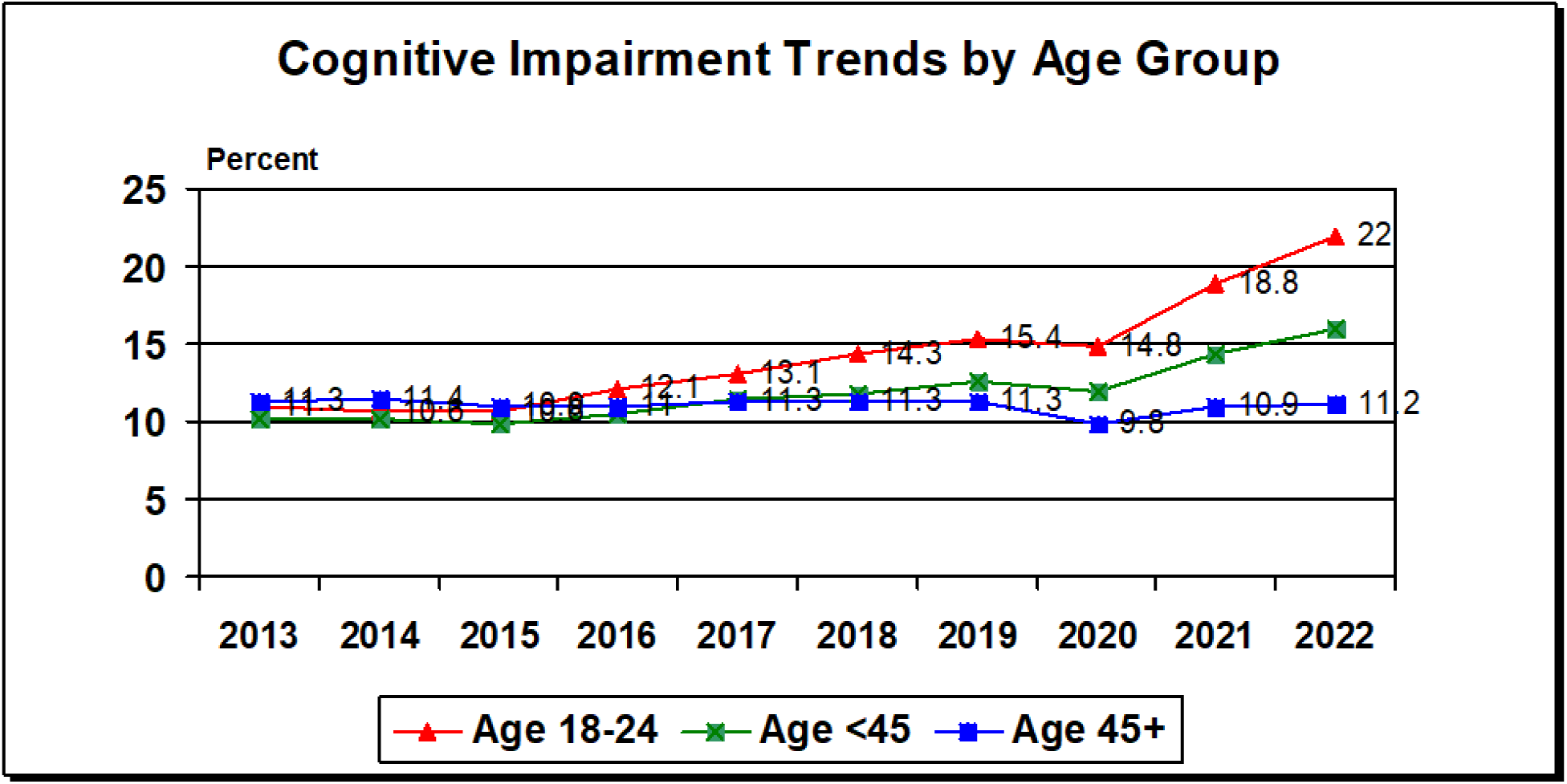
Cognitive Impairment (CI) Trends by Age Group, 2013-2022 Behavioral Risk Factor Surveillance System. CI: Because of a physical, mental, or emotional condition, do you have serious difficulty concentrating, remembering, or making decisions?

**Table 1.**
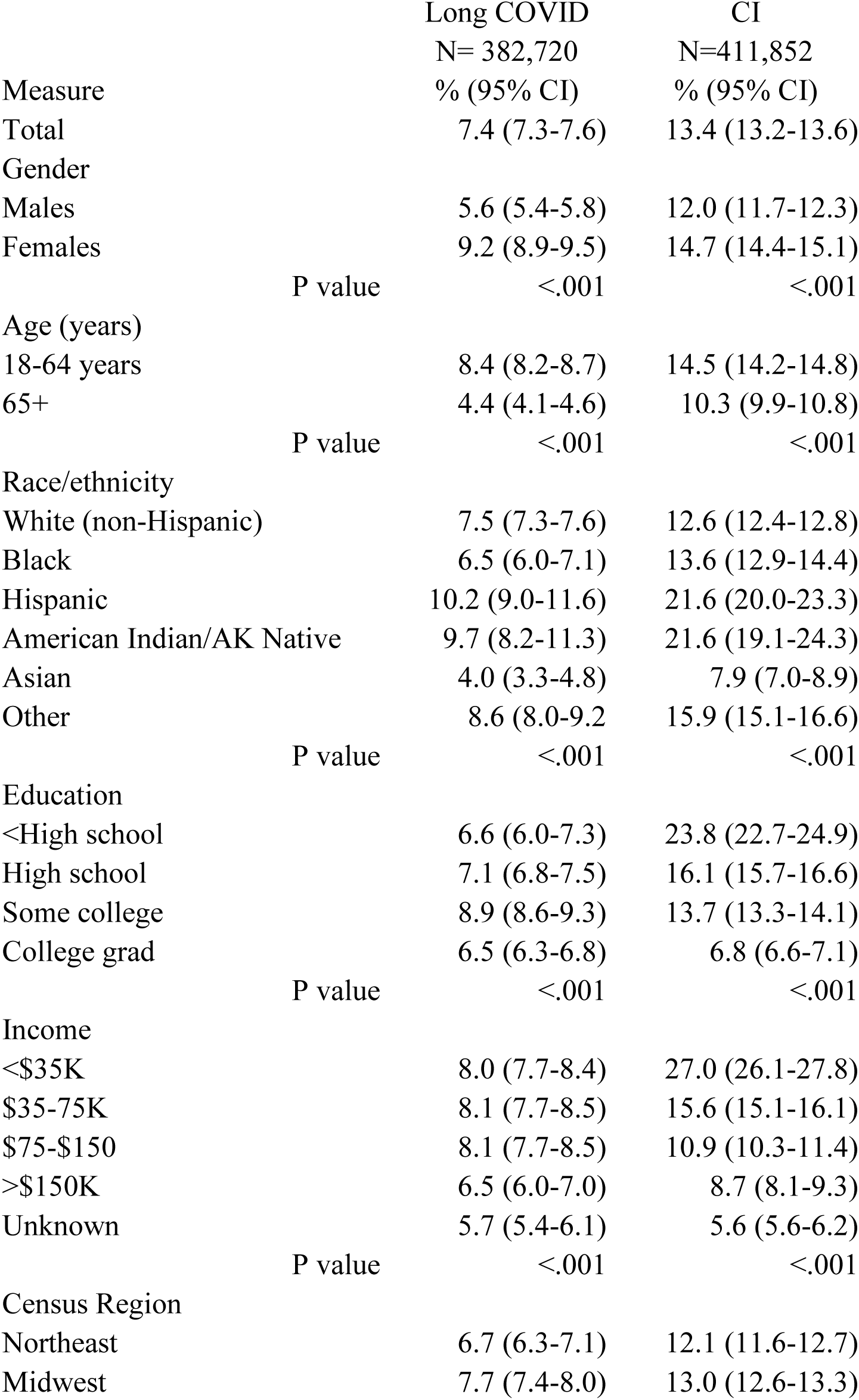
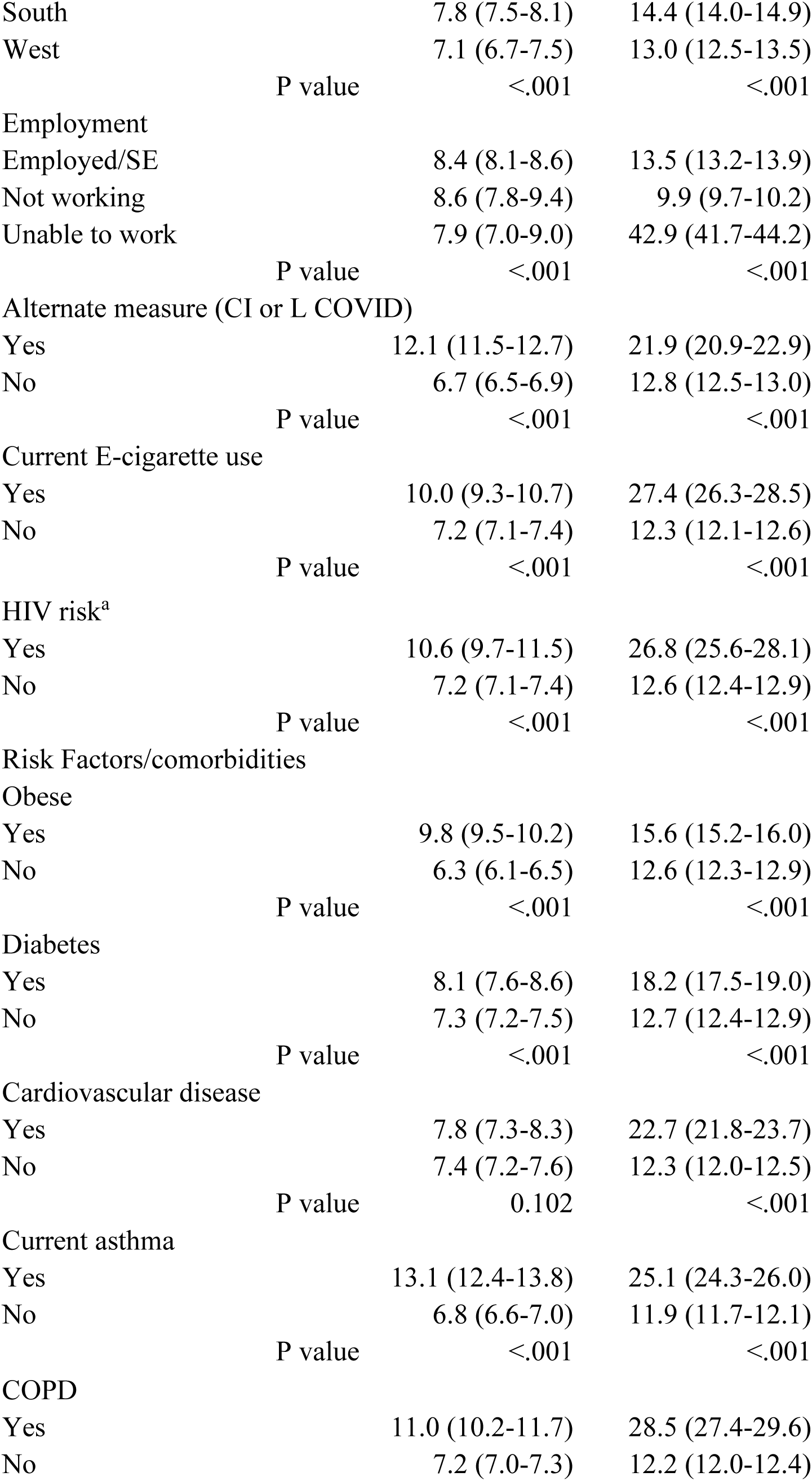
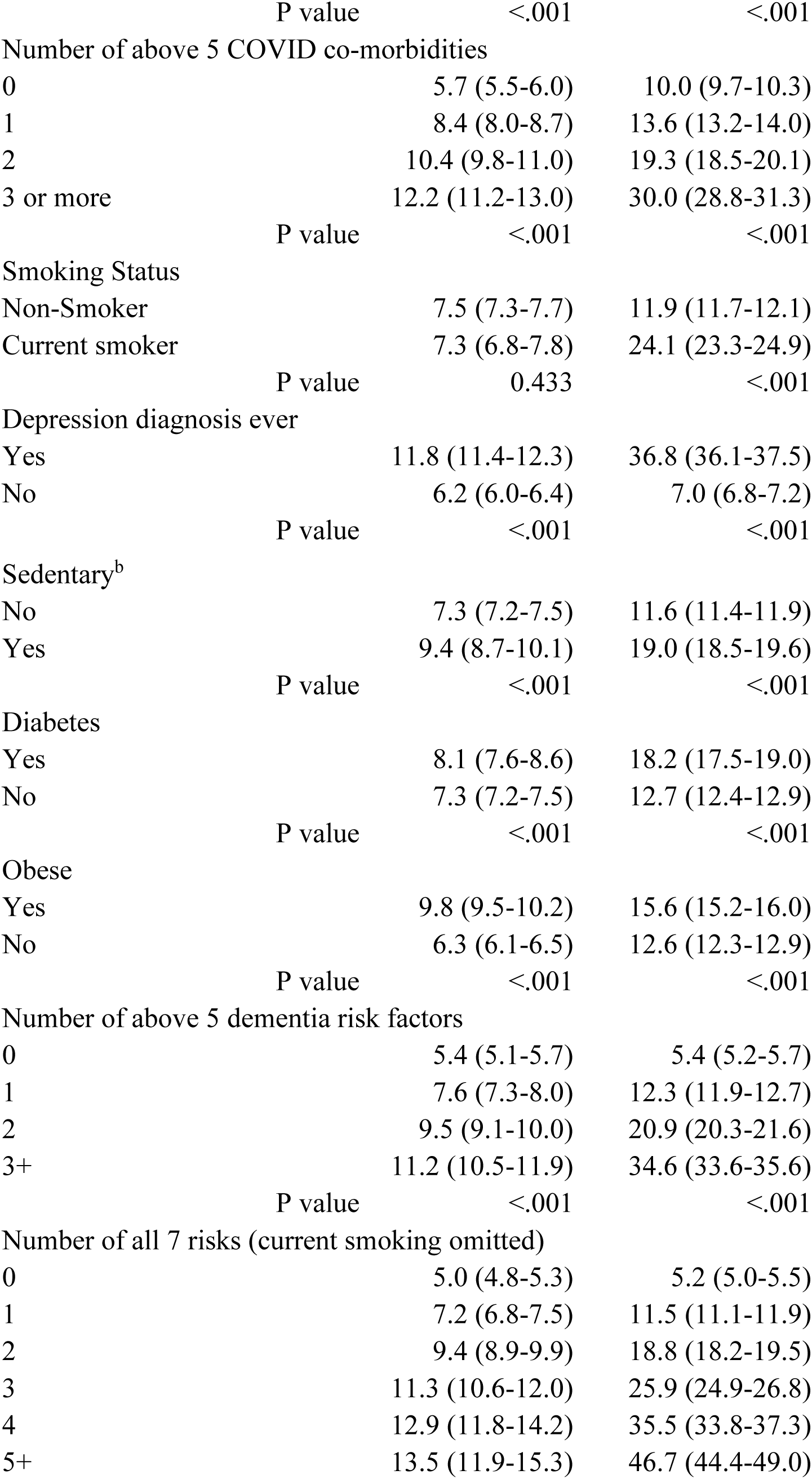
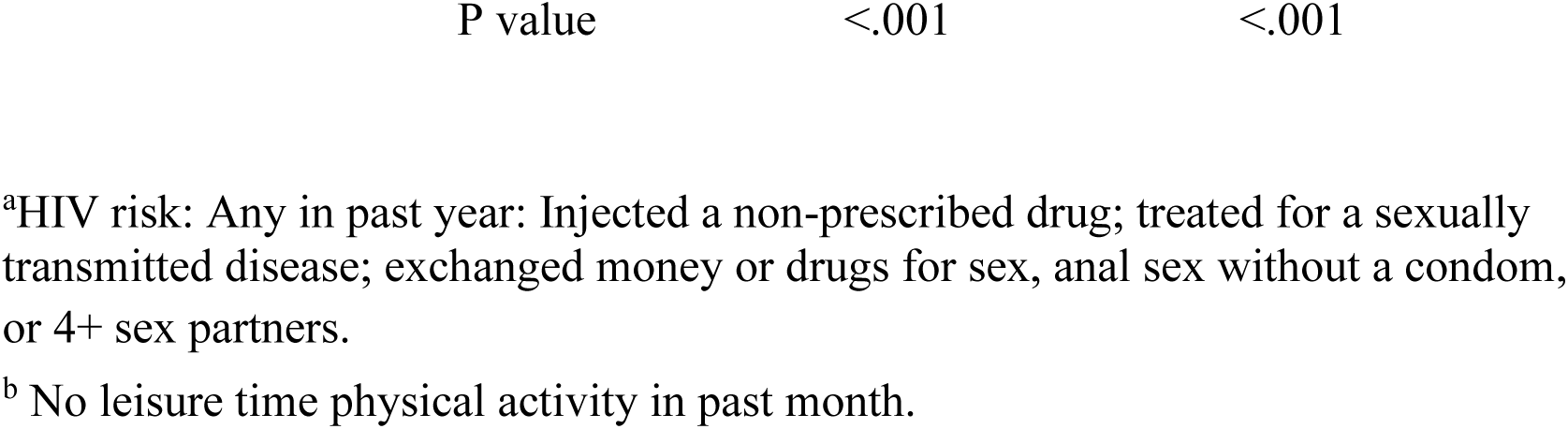
Percentages and 95% CIs for adults with Long COVID and Cognitive Impairment (CI); Results from weighted analysis in Stata, 2022 Behavioral Risk Factor Surveillance System, 50 states & DC.

Results of logistic regression for long COVID and CI (Table 2), confirmed most unadjusted results in Table 1 except sedentary lifestyle was no longer significant for long COVID and obesity was no longer significant for CI. Adjusted results found that the highest adjusted odds ratios (AORs) for both measures were for 5 or more of the 7 risk factors combining COVID and dementia risks, with AOR= 3.01 for long COVID and 10.5 for CI.

**Table 2.**
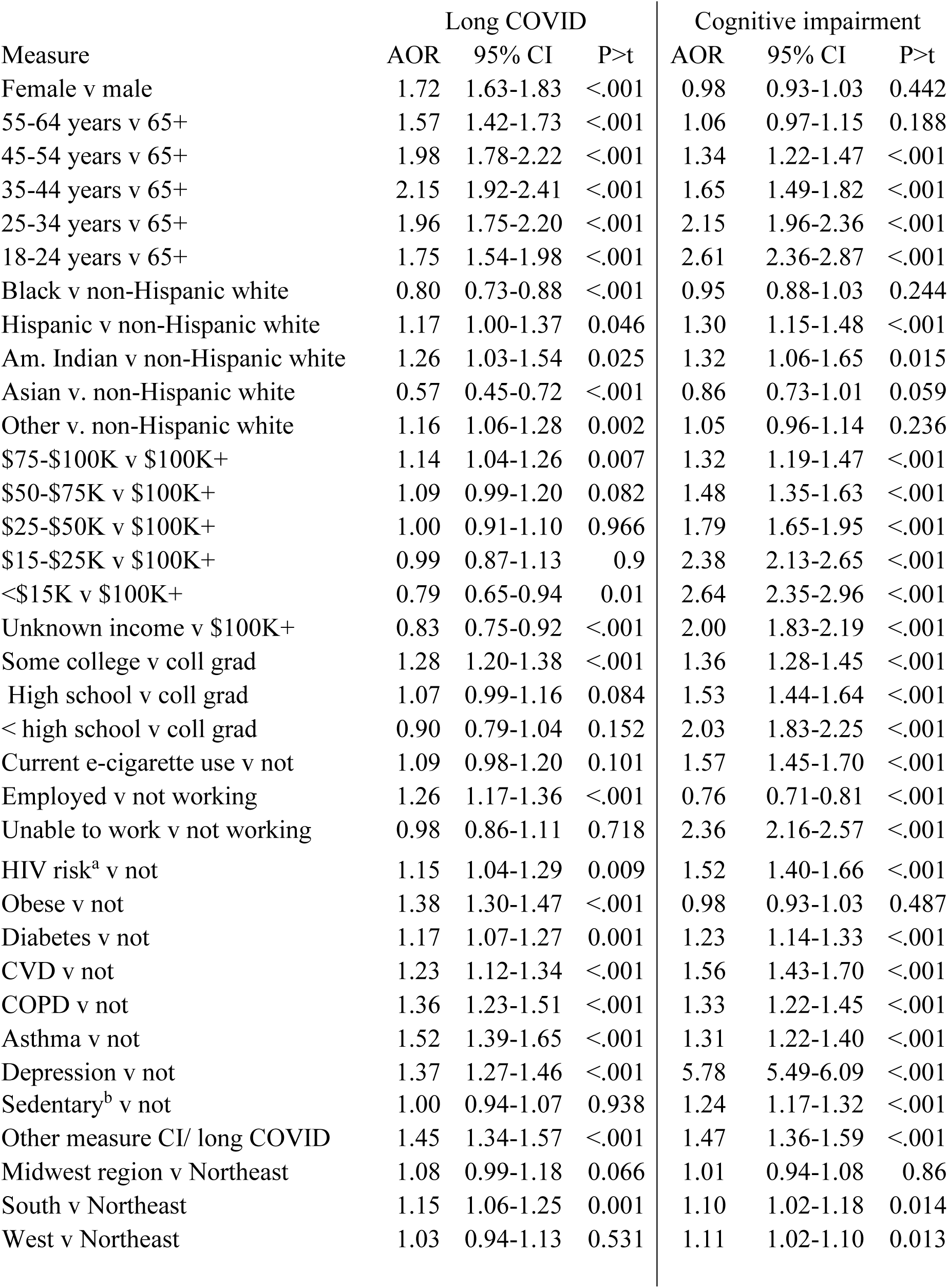
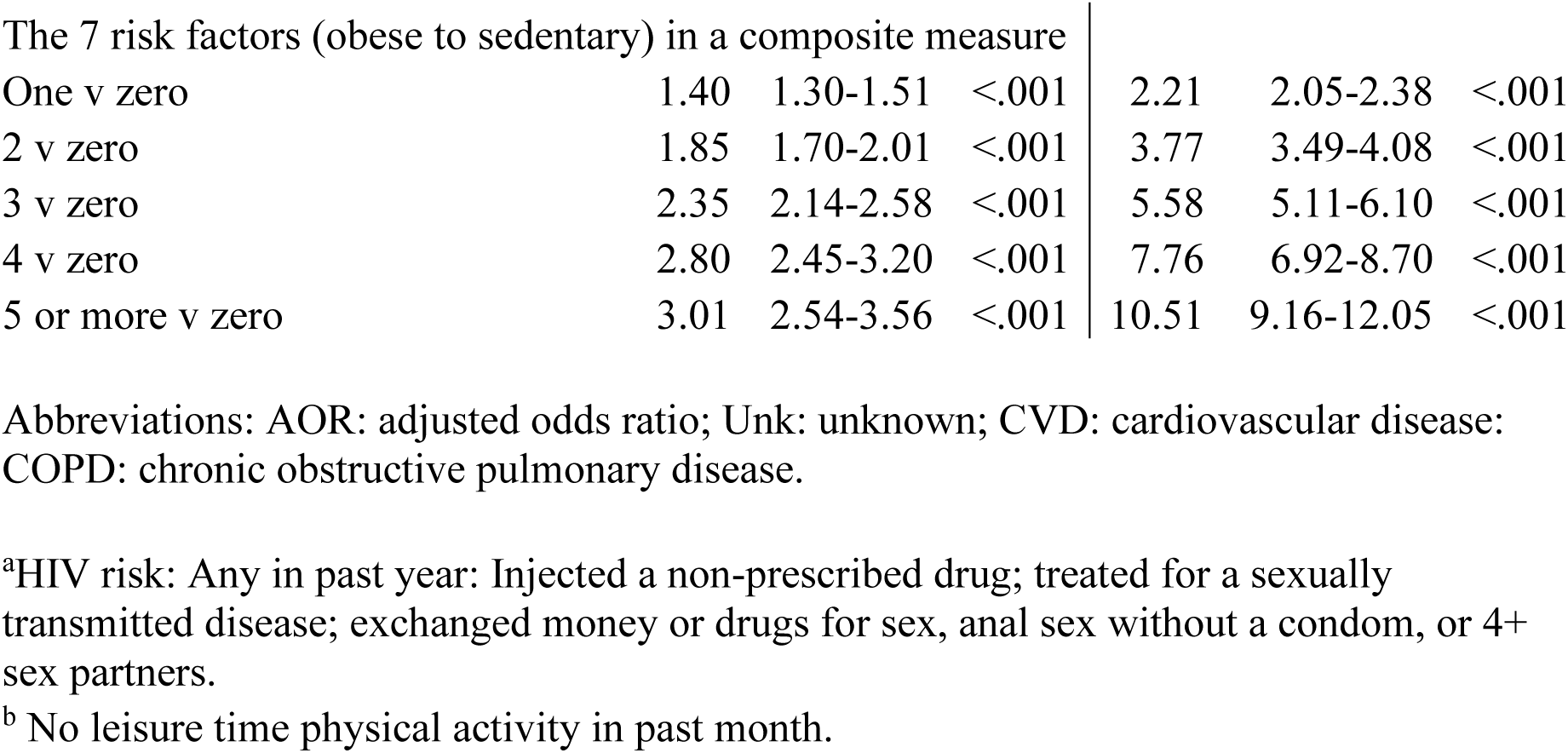
Results of logistic regression with all measures shown included in the model; 2022 Behavioral Risk Factor Surveillance System. N=329,127.

| Measure | Long COVID |  |  | Cognitive impairment |  |  |
| --- | --- | --- | --- | --- | --- | --- |
|  | AOR | 95% CI | P>t | AOR | 95% CI | P>t |
| Female v male | 1.72 | 1.63-1.83 | <.001 | 0.98 | 0.93-1.03 | 0.442 |
| 55-64 years v 65+ | 1.57 | 1.42-1.73 | <.001 | 1.06 | 0.97-1.15 | 0.188 |
| 45-54 years v 65+ | 1.98 | 1.78-2.22 | <.001 | 1.34 | 1.22-1.47 | <.001 |
| 35-44 years v 65+ | 2.15 | 1.92-2.41 | <.001 | 1.65 | 1.49-1.82 | <.001 |
| 25-34 years v 65+ | 1.96 | 1.75-2.20 | <.001 | 2.15 | 1.96-2.36 | <.001 |
| 18-24 years v 65+ | 1.75 | 1.54-1.98 | <.001 | 2.61 | 2.36-2.87 | <.001 |
| Black v non-Hispanic white | 0.80 | 0.73-0.88 | <.001 | 0.95 | 0.88-1.03 | 0.244 |
| Hispanic v non-Hispanic white | 1.17 | 1.00-1.37 | 0.046 | 1.30 | 1.15-1.48 | <.001 |
| Am. Indian v non-Hispanic white | 1.26 | 1.03-1.54 | 0.025 | 1.32 | 1.06-1.65 | 0.015 |
| Asian v. non-Hispanic white | 0.57 | 0.45-0.72 | <.001 | 0.86 | 0.73-1.01 | 0.059 |
| Other v. non-Hispanic white | 1.16 | 1.06-1.28 | 0.002 | 1.05 | 0.96-1.14 | 0.236 |
| \$75-\$100K v \$100K+ | 1.14 | 1.04-1.26 | 0.007 | 1.32 | 1.19-1.47 | <.001 |
| \$50-\$75K v \$100K+ | 1.09 | 0.99-1.20 | 0.082 | 1.48 | 1.35-1.63 | <.001 |
| \$25-\$50K v \$100K+ | 1.00 | 0.91-1.10 | 0.966 | 1.79 | 1.65-1.95 | <.001 |
| \$15-\$25K v \$100K+ | 0.99 | 0.87-1.13 | 0.9 | 2.38 | 2.13-2.65 | <.001 |
| <\$15K v \$100K+ | 0.79 | 0.65-0.94 | 0.01 | 2.64 | 2.35-2.96 | <.001 |
| Unknown income v \$100K+ | 0.83 | 0.75-0.92 | <.001 | 2.00 | 1.83-2.19 | <.001 |
| Some college v coll grad | 1.28 | 1.20-1.38 | <.001 | 1.36 | 1.28-1.45 | <.001 |
| High school v coll grad | 1.07 | 0.99-1.16 | 0.084 | 1.53 | 1.44-1.64 | <.001 |
| < high school v coll grad | 0.90 | 0.79-1.04 | 0.152 | 2.03 | 1.83-2.25 | <.001 |
| Current e-cigarette use v not | 1.09 | 0.98-1.20 | 0.101 | 1.57 | 1.45-1.70 | <.001 |
| Employed v not working | 1.26 | 1.17-1.36 | <.001 | 0.76 | 0.71-0.81 | <.001 |
| Unable to work v not working | 0.98 | 0.86-1.11 | 0.718 | 2.36 | 2.16-2.57 | <.001 |
| HIV risk<sup>a</sup> v not | 1.15 | 1.04-1.29 | 0.009 | 1.52 | 1.40-1.66 | <.001 |
| Obese v not | 1.38 | 1.30-1.47 | <.001 | 0.98 | 0.93-1.03 | 0.487 |
| Diabetes v not | 1.17 | 1.07-1.27 | 0.001 | 1.23 | 1.14-1.33 | <.001 |
| CVD v not | 1.23 | 1.12-1.34 | <.001 | 1.56 | 1.43-1.70 | <.001 |
| COPD v not | 1.36 | 1.23-1.51 | <.001 | 1.33 | 1.22-1.45 | <.001 |
| Asthma v not | 1.52 | 1.39-1.65 | <.001 | 1.31 | 1.22-1.40 | <.001 |
| Depression v not | 1.37 | 1.27-1.46 | <.001 | 5.78 | 5.49-6.09 | <.001 |
| Sedentary<sup>b</sup> v not | 1.00 | 0.94-1.07 | 0.938 | 1.24 | 1.17-1.32 | <.001 |
| Other measure CI/ long COVID | 1.45 | 1.34-1.57 | <.001 | 1.47 | 1.36-1.59 | <.001 |
| Midwest region v Northeast | 1.08 | 0.99-1.18 | 0.066 | 1.01 | 0.94-1.08 | 0.86 |
| South v Northeast | 1.15 | 1.06-1.25 | 0.001 | 1.10 | 1.02-1.18 | 0.014 |
| West v Northeast | 1.03 | 0.94-1.13 | 0.531 | 1.11 | 1.02-1.10 | 0.013 |
| The 7 risk factors (obese to sedentary) in a composite measure |  |  |  |  |  |  |
| One v zero | 1.40 | 1.30-1.51 | <.001 | 2.21 | 2.05-2.38 | <.001 |
| 2 v zero | 1.85 | 1.70-2.01 | <.001 | 3.77 | 3.49-4.08 | <.001 |
| 3 v zero | 2.35 | 2.14-2.58 | <.001 | 5.58 | 5.11-6.10 | <.001 |
| 4 v zero | 2.80 | 2.45-3.20 | <.001 | 7.76 | 6.92-8.70 | <.001 |
| 5 or more v zero | 3.01 | 2.54-3.56 | <.001 | 10.51 | 9.16-12.05 | <.001 |

Abbreviations: AOR: adjusted odds ratio; Unk: unknown; CVD: cardiovascular disease; COPD: chronic obstructive pulmonary disease.
<sup>a</sup>HIV risk: Any in past year: Injected a non-prescribed drug; treated for a sexually transmitted disease; exchanged money or drugs for sex, anal sex without a condom, or 4+ sex partners.
<sup>b</sup> No leisure time physical activity in past month.

Vaccination rates for ≥3 doses were 46.9% overall and varied by age, COVID, and CI status. Lowest vaccination rates (40% or lower) were found for those 18-64 years, and those with either long COVID or CI, and highest rates for those ages ≥65 years at 67.8%. High vaccination rates (>50%) were also reported for adults with diabetes and CVD, consistent with influenza vaccination rates (not shown). Apparent vaccine effectiveness of ≥3 doses vs <3 was 38.0% for long COVID, reducing rates from 9.2% for <3 doses to 5.7% for ≥3 (12), and 35.2% for CI, reducing CI rates from 15.9% to 10.3% (Table 3). Rates for adults with ≥3 doses were reduced to those comparable to adults with 0 of the 5 COVID risks (5.7 for long COVID and 10.0% for CI as shown in Table 1) but not down to rates for 0 dementia risks which were 5.4% for both long COVID and CI. Results for CI limited to respondents without long COVID or who never tested positive for COVID (Table 3) were similar to results for all respondents with CI (34.4% - 36.9% vs.35.1% for all with CI). When limited to adults **with** long COVID, prevalence rates were higher for both vaccine doses and the apparent effectiveness was lower. Dose response gradients were shown for 1 to 4+ vaccine doses vs 0 for both outcomes, also shown in Table 3. Vaccines were apparently not effective for either outcome for ages 18-24 years, with P values >0.05 and negative estimated effectiveness (not shown). For comparison, apparent vaccine effectiveness for the 34.8% of all adults with a positive COVID test was 21.4%, 16.2% for those with a positive test and not long COVID, and for COPD, asthma, obesity, and depression vaccine effectiveness was approximately 5% or less. For CVD and diabetes, apparently due to high vaccination rates for adults with those conditions, the apparent effectiveness of vaccines was <u>negative</u> 33% or 49%. Logistic regression that included age in 6 groups, gender, region, the alternate measure (CI or long COVID) and the measure of ≥3 vs < 3 vaccines confirmed apparent effectiveness with AOR=0.66 for the vaccine measure for long COVID and AOR= 0.70 for CI.

**Table 3.** Rates of long COVID and cognitive impairment at various doses of COVID vaccine with % reduction representing apparent effectiveness. 2022 Behavioral Risk Factor Surveillance System, 29 states^a^, N=168,822.

| **Ages** | **Long COVID** |  |  | **Cognitive impairment** |  |  |
| --- | --- | --- | --- | --- | --- | --- |
|  | **Prevalence rates** |  |  | **Prevalence rates** |  |  |
|  | **<3 vax** | **≥3 vax** | **% Reduction** | **<3 vax** | **≥3 vax** | **% Reduction** |
| All ages | 9.2 | 5.7 | 38.0% | 15.9 | 10.3 | 35.2% |
| 65+ | 5.8 | 3.9 | 32.8% | 13.2 | 8.8 | 33.3% |
| 45+ | 9.0 | 5.2 | 42.2% | 13.8 | 8.9 | 35.5% |
| <45 | 9.8 | 7.2 | 26.5% | 17.8 | 13.8 | 22.5% |
| All ages | N=115,011 never tested positive |  |  | 16.0 | 10.1 | 36.9% |
| All ages | N=155,505 without long COVID |  |  | 15.1 | 9.9 | 34.4% |
| All ages | Limited to 4,037 w/ long COVID |  |  | 23.9 | 16.7 | 30.1% |

Dose-response results for all ages, # vaccine doses vs 0, N varies.
| **# doses** | **Long COVID** |  |  | **Cognitive impairment** |  |  |
| --- | --- | --- | --- | --- | --- | --- |
|  | **0 doses** | **Max dose** | **% Reduction** | **0 doses** | **Max dose** | **% Reduction** |
| 1 | 9.5 | 9.9 | ---- | 16.7 | 18.2 | ---- |
| 2 | 9.5 | 8.9 | 6.3% | 16.7 | 14.9 | 10.8% |
| 3 | 9.5 | 5.9 | 37.9% | 16.7 | 10.8 | 35.3% |
| 4 + | 9.5 | 5.0 | 47.4% | 16.7 | 8.7 | 47.9% |

Long COVID: symptoms lasting ≥3 months.
Cognitive impairment: serious difficulty concentrating, remembering, or making decisions, question asked on all federal surveys.
<sup>a</sup> AR, CT, DE, GA, HI, ID, IL, IA, KS, LA, ME, MD, MA, MT, NE, NH, NJ, NM, NY, NC, ND, OK, RI, SC, TN, TX, WV, WI, WY

## Discussion

Please bear with us as we try to make sense of results showing that the apparent effectiveness of COVID vaccines on cognitive impairment (35.2%) is greater than the effectiveness on COVID itself as measured by a positive test (21.4%). Study results show that CI and long COVID are indeed related, but they also raise many questions. Starting with Figure 1, the trend line for CI for ages 18-44 appears to suggest an association with COVID, as the slope of the line for adults younger than age 45 years seems to increase when the pandemic started in 2020. That could be a coincidence and there could be several factors causing CI rates to increase after 2020 other than the pandemic. But it is an additional piece of information to keep in mind indicative of COVD and CI being related.

Results in Tables 1 and 2 both indicate that having the alternate measure (CI for long COVID and vice versa) increases the prevalence rate and when controlled for all the other measures in the logistic regression model, the AOR is 1.45 for CI in the long COVID model or 1.47 for long COVID in the CI model. Results for the various risk factors appear similar for long COVID and CI, with the observation that all three composite risk factor measures – for COVID risks, dementia risks, and both combined - appear to have greater dose response gradients for CI compared with long COVID but all show some degree of dose-response. This is most obvious in the logistic regression results in Table 2 where the AOR for 5 or more of the 7 combined risk factors is 10.51 for CI and 3.01 for long COVID. Clearly, most of the risk factors, separately or in groups, increase rates of both outcomes. More information comes from a study using these same BRFSS data (12) and this same measure for long COVID, compared with adults who had a positive test that did not develop long COVID (termed “Just COVID”). This latter group represented 4 in 5 adults with a positive test and 26.5% of all adults. Those with Just COVID reported the most favorable responses for 15 of 17 measures of health and disability status while those with long COVID reported worse results for 13 of the 17. Contrary to results in this paper (Table 1) showing that rates of both long COVID and CI increased at least 2-fold between 0 and 3 or more COVID morbidities, for Just COVID, rates were highest for those with 0 COVID comorbidities (27.7%) and decreased to 22.0% for those with 3 or more of the 5 risks (12). This suggests a COVID measure that is very different from long COVID (and CI), having different risk factors and less negative impact on health status.

Another study provides a direct comparison of adults ages 45 and older with CI only, subjective cognitive decline (SCD) only, and both (13). That study used 2015 BRFSS data which included 35 states that asked the cognitive decline module and used different terminology. The study used the same composite measure of dementia risk as used in this study but included hypertension which is missing here. Approximately 11% of adults were in each of the groups of only SCD, only CI, and the combination, with about half of those with each separate measure also reporting the other. Results showed that in general, there was a progression of adverse effects from SCD only, CI only, to adults with both. Logistic regression results for the composite measure of dementia risk found the highest AORs for those with all 6 risk factors (obesity, diabetes, being sedentary, hypertension, smoking, depression) of 7.6 for SCD and 24.5 for CI. These results suggest that for adults ages 45+, cognitive decline is less affected by the 6 risk factors than CI and produces a smaller adverse effect compared with CI.

In terms of prevention strategies, the effects shown in Table 3 for ≥3 COVID vaccines indicate that long COVID rates drop to 5.7% and CI rates drop to 10.3%. To achieve a comparable drop in rates using standard chronic disease strategies of reducing risk factors would require total elimination of the 5 COVID risks and dementia risks and the total number of combined risks would need to be reduced to between 0 and 1 risk factor (see Table 1). Neither option would be easy, but knowing how hard reducing the obesity rate is (14), which is related to many of these risk factors, vaccination might provide a better choice.

Other studies adding to the body of knowledge on long COVID, CI, and vaccines include one that first confirmed vaccines reduced risk of long COVID (15). That study confirmed our results that COVID vaccines reduce the risk of developing long COVID compared with those not vaccinated, and also that vaccination reduced the risk of cognitive impairment (as a symptom) in those with long COVID. This latter finding is not directly comparable to our findings because we were able to show vaccine effectiveness for CI among people **without** long COVID or a positive COVID test to assure we were not measuring any vaccine effect due to the presence of COVID.

This might be a good time to address how mRNA vaccines work. The COVID mRNA vaccine uses the SARS-CoV-2 spike protein as antigen to create an immune response without exposing the vaccine recipient to the virus itself – just the synthetic mRNA that makes the spike protein (16). Thus, the immune response created is against a protein – the spike protein on the virus.

When the vaccine recipient it exposed to the COVID virus, their immune system will recognize it and attack the virus. Because COVID (and the SARs virus) and CI share so many characteristics and risk factors, it seems at least plausible that the immune system of a COVID vaccine recipient might also recognize and attack a protein similar to the spike protein.

One of the key features of Alzheimer’s Disease (AD) is amyloid plaques in the brain composed of amyloid-β (Aβ) proteins, which can form as early as 20 years prior to clinical symptoms (17).

In a key study (18) an amyloid precursor protein (APP), precursor of the Aβ proteins of AD, was found to interact with the spike protein of SARS-CoV-2, the protein the COVID vaccine was designed to attack. Those findings seem to help fill in gaps in understanding the similarities between AD and COVID-19 and add to the plausibility for our vaccine results on CI.

Is COVID-19 a risk factor for AD? In one large study of adults ages ≥65 years (19), those with COVID-19 were at significantly increased risk for AD after a COVID-19 diagnosis (hazard ratio or HR:1.69, 95% CI: 1.53-1.72). Risk was highest in adults age ≥85 years and in women.

Another study noted that survivors of COVID appear at increased risk of developing AD (17) suggesting COVID-19 will remain a threat to our healthcare systems due to AD cases for the foreseeable future. On the other hand, at least one study found evidence that AD was a risk factor for COVID (19). Their study found that patients with AD were more susceptible to COVID and more likely to report adverse clinical outcomes compared with patients without AD.

When considering strategies for prevention of COVID and related conditions there are several notable findings from this study. One important observation seems to be that vaccines were not very effective for adults ages 18-24 years. The difference seen between CI trends for those 45 and older and <45years are also worthy of comment especially when considering results (13) showing that adults with CI appear to have more adverse effects compared with adults with cognitive decline where data are lacking for those < 45 years. Both vaccines and risk factors appear to be key to reducing the effects of long COVID, CI, and likely cognitive decline and dementia too, but not for adults who have a positive COVID test who don’t develop long COVID. Those adults seem to be less adversely impacted by COVID but could still infect others. These results for adults with just a positive COVID test show that they can be distinguished from those who develop long COVID by their relative lack of risk factors (obesity, diabetes, hypertension, sedentary lifestyle, depression, COPD, CVD, and asthma) and to the apparent low effectiveness of COVID vaccines on them. Our key finding is that COVID vaccines appear as effective against CI as they are against long COVID which is more effective than they are against a positive COVID test.

Limitations: There are at least six limitations to this study. First, because the BRFSS only surveys households, among the institutions the survey omits are nursing homes and prisons, which appeared to have high rates of COVID especially early in the pandemic. Thus, results may underestimate the true rates of CI and long COVID. Second, results are self-reported and except as noted for COVID are not based on an actual test or diagnosis. Third, because BRFSS is a telephone survey, only respondents able to complete a survey over the telephone are included. Another BRFSS study that included non-respondents in households with respondents found that some measures of cognitive decline were under-reported by as much as 70% when only respondent data was included (21). Fourth, the lack of a measure of hypertension on the survey for 2022 meant that the composite measure of COVID comorbidities lacked a key component (10). Fifth, survey results can’t distinguish cause and effect. Sixth, only 29 states had survey data on COVID vaccines.

## Conclusion

Although our study leaves many questions unanswered – such as HOW the vaccines work and if they can slow progression to AD - that does not mean the results aren’t valid or useful. Nothing in the cited studies appears to question the plausibility of our results. On the contrary, findings confirm many similarities between COVID and CI (or AD) and suggest that a precursor protein associated with AD might be similar enough to the spike protein targeted by the vaccines to stimulate an immune response. It should not be difficult to confirm these results with data that include vaccines received and a measure of cognitive impairment and to assure that the results are not compromised by COVID. The opportunities these results suggest for reducing cognitive impairment, whether it progresses to AD or not, are exciting.

## Data Availability

Data available at: Behavioral Risk Factor Surveillance System (BRFSS). Centers for Disease Control and Prevention. Behavioral Risk Factor Surveillance System. Annual Survey Data. https://www.cdc.gov/brfss/annual_data/annual_data.htm. Accessed September 1, 2023.

## Acknowledgment

This study was partially supported by a cooperative agreement from CDC (#1NU58DP006901) which had no role in the writing of the manuscript.

